# Large-scale alternative polyadenylation (APA)-wide association studies to identify putative susceptibility genes in human common cancers

**DOI:** 10.1101/2023.11.05.23298125

**Authors:** Xingyi Guo, Jie Ping, Yaohua Yang, Xinwan Su, Xiao-ou Shu, Wanqing Wen, Zhishan Chen, Yunjing Zhang, Ran Tao, Guochong Jia, Jingni He, Qiuyin Cai, Qingrun Zhang, Graham G Giles, Rachel Pearlman, Gad Rennert, Pavel Vodicka, Amanda Phipps, Stephen B Gruber, Graham Casey, Ulrike Peters, Jirong Long, Weiqiang Lin, Wei Zheng

## Abstract

Alternative polyadenylation (APA) modulates mRNA processing in the 3’ untranslated regions (3’UTR), which affect mRNA stability and translation efficiency. Here, we build genetic models to predict APA levels in multiple tissues using sequencing data of 1,337 samples from the Genotype-Tissue Expression, and apply these models to assess associations between genetically predicted APA levels and cancer risk with data from large genome-wide association studies of six common cancers, including breast, ovary, prostate, colorectum, lung, and pancreas among European-ancestry populations. At a Bonferroni-corrected *P*□<□0.05, we identify 58 risk genes, including seven in newly identified loci. Using luciferase reporter assays, we demonstrate that risk alleles of 3’UTR variants, rs324015 (*STAT6*), rs2280503 (*DIP2B*), rs1128450 (*FBXO38*) and rs145220637 (*LDAH*), could significantly increase post-transcriptional activities of their target genes compared to reference alleles. Further gene knockdown experiments confirm their oncogenic roles. Our study provides additional insight into the genetic susceptibility of these common cancers.

## Introduction

Genome-wide association studies (GWAS) have identified common variants in approximately 1000 genetic loci associated with risk of human cancers^1, 2, 3, 4, 5^. Nearly 90% of GWAS-identified risk variants are located in non-coding or intergenic regions, suggesting that their associations with cancer risk may be mediated through regulatory roles in gene expression. Significant efforts have been made to identify potential target genes and biological mechanisms driving cancer susceptibility in these GWAS-identified risk loci. Previous expression quantitative trait loci (eQTL) analyses, including our own^6, 7^, have discovered several putative susceptibility genes in human cancers, primarily based on RNA sequencing (RNA-seq) data in target tissues^1, 7, 8, 9^. Since 2015, multiple transcriptome-wide association studies (TWAS) have been conducted to investigate associations between genetically predicted gene expression and cancer risk^10, 11, 12, 13, 14, 15, 16, 17^. Unlike conventional eQTL analyses and GWAS, TWAS use aggregated information from multiple *cis*-genetic variants, thus may have higher statistical power to identify novel association signals overlooked in GWAS^18^. We and others have identified large numbers of putative susceptibility genes through TWAS for several cancers including breast, ovarian, colorectal, pancreatic, and prostate cancers^12, 13, 14, 15, 16, 17^. However, TWAS primarily focus on eQTLs. Genetic variants related to posttranscriptional regulation, such as the 3’ untranslated region (UTR) alternative polyadenylation (APA) QTL (3’aQTLs), are largely unexplored in association studies with cancer risk.

APA modulates mRNA processing at different polyadenylation (poly[A]) sites and thus affects the length of 3’ untranslated regions (UTR), which influences *cis-*regulatory elements, such as microRNAs (miRNA) or RNA-binding protein (RBP) binding sites^19^. Therefore, APA can affect mRNA stability and translation efficiency^20^. Recent studies suggest that 3’aQTLs account for a significant proportion of APA, subsequently affecting disease heritability^21, 22, 23^. Therefore, conducting alternative polyadenylation-wide association studies (APA-WAS) to investigate associations of genetically predicted APA levels with cancer risk may help identify novel risk loci and putative susceptibility genes for cancer and improve the biological understanding of cancer susceptibility. In this study, we used RNA-sequencing (RNA-seq) data generated in multiple normal tissues, along with the matched whole genome sequencing (WGS) data generated in blood samples from the Genotype-Tissue Expression (GTEx), and large-scale GWAS data for cancers of breast, ovary, prostate, colorectum, lung, and pancreas to conduct APA-WAS to search for susceptibility genes and loci of these common cancers.

## Results

### APA levels predicted using *cis*-genetic variants in normal breast, ovarian, prostate, colon transverse, lung, and pancreas tissues

We utilized DaPars v.2 regression framework^21^ to identify APA events by analyzing RNA-seq data generated in normal tissues of breast (n=114), ovary (n=127), prostate (n=170), colon transverse (n=285), lung (n=410), and pancreas (n=231) obtained from individuals of European descendants included in the GTEx (see Methods; Figure 1A). These APA levels were quantified using the percentage of distal polyA site usage index (PDUI) of each 3′UTR APA. Numbers of detected APA events identified in our study were 23,502 for breast (13,276 genes), 21,995 for ovary (12,384 genes), 23,498 for prostate (13,337 genes), 23,099 for colon transverse (13,152 genes), 23,448 for lung (13,221 genes), and 19,080 for pancreas (10,989 genes), which is comparable to those reported from a previous study^21^. We then used genotype data and PDUI of these APA events to build models to predict APA levels (measured PDUI) using *cis*- genetic variants (flanking ± 500Kb region) with the elastic-net approach^10^ for each specific tissue (Figure 1B). Using the approach similar to that described in our previous TWAS^12,16^, we conducted association analyses with cancer risk using models that predicted APA levels at a prediction performance of R^2^ > 0.01 at *P* < 0.05, which included 2,556 models for breast, 2,363 models for ovary, 2,631 models for prostate, 2,913 models for colon transverse, 3,098 models for lung, and 1,736 models for pancreas (Table S1).

**Figure 1.**
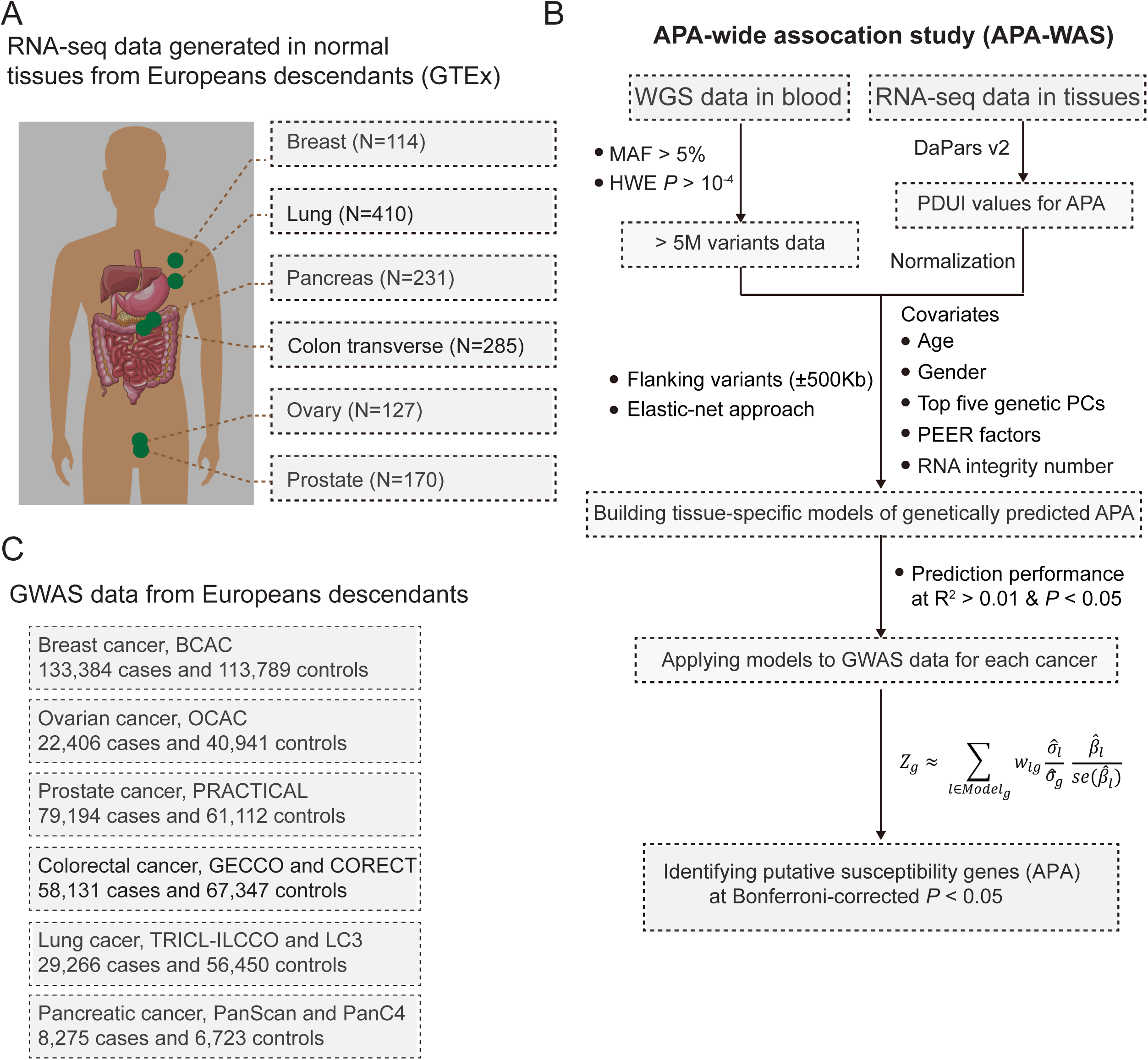
Study design of APA-WAS in cancers of breast, ovary, prostate, colorectum, lung, and pancreas. A) Each box shows the reference data used to build models for breast, ovarian, pancreas, colorectal, prostate, and lung cancers. B) A workflow to illustrate our analytic framework, including model building for genetically predicted APA levels and association analyses to identify putative cancer susceptibility genes. C) Each box shows the reference GWAS data for each APA-WAS.

### Identification of putative susceptibility genes for cancers of breast, ovary, prostate, colorectum, lung, and pancreas

To evaluate associations of genetically predicted APA levels with cancer risk, we applied the above prediction models to the GWAS summary statistic data from cancers of breast (N=247,173), ovary (N=63,347), prostate (N=140,306), colorectum (N=125,478), lung (N=85,716), and pancreas (N=21,536) (Figure 1C, Table S2, see Method). We identified putative susceptibility genes for each cancer using a Bonferroni-corrected significance level at *P* < 0.05 (see **Methods**). To further investigate whether the identified associations were independent of the established GWAS-identified associations, we conducted conditional APA-WAS analysis by adjusting for the nearest GWAS-identified risk variants (index variants; see **Methods**). Furthermore, we comprehensively compared our findings with those identified by eQTL analysis in GTEx and reported from previous TWAS, eQTL, and fine-mapping studies for breast ^1, 7, 14, 24, 25, 26^, ovary^6, 16^, prostate^5, 6, 13, 26, 27^, colorectal^12, 28, 29^, and lung cancers^6, 26, 30, 31^.

For breast cancer, our analysis revealed 18 significant APA events, corresponding to 14 putative susceptibility genes (Figure 2; Table S3). Of them, eight (*AMFR, RPS23, ARL17A, ARL18A, COX11, NSUN4*, *P4HA2,* and *STAG3L2*) were reported in previous genetic studies (Table S4). Of the remaining six newly identified putative susceptibility genes, three are located at loci more than 500Kb away from any GWAS-identified breast cancer risk variants, including *KDSR* (18q21.33), *PML* (15q24.1), and *SSR2* (1q22) (Table 1). Conditional analyses showed that all three genes remained statistically significant at a Bonferroni-corrected threshold of *P* < 0.0028 (0.05/18 tests), while none of the other three putative susceptibility genes (*SARAF, ANO8,* and *SDHA*) located in GWAS-identified risk loci showed a significant association after adjusting for the nearest index variants (Table 1). Importantly, it should be highlighted that *SSR2* exhibits a partial correlation with known GWAS variants, albeit with marginal significance. Therefore, it would be advisable to gather more compelling and convincing data to verifying the finding. For ovarian cancer, we identified two previously reported genes (*RCCD1*) at 15q26.1 and (*ARL17B*) at 17q21.31 (Figure 2; Table S3 and S5). Specifically, *ARL17B is* located 839kb away from the nearest index variant, rs1879586^16^. The association of ovarian cancer risk with this gene has been reported in previous TWAS in ovarian cancer^17^. For prostate cancer, we identified 38 significant APA events, corresponding to 27 putative susceptibility genes (Figure 2; Table S3). Of them, 15 genes were reported in previous studies (Table S6). Conditional analysis showed that eight of the remaining 12 newly identified genes (*FBXO38, STMN3, MYC, ADGRG1, THADA, PPP2R2A, WASHC2C*, and *ST3GAL5*) remained statistically significant at Bonferroni-corrected *P* < 0.05, including one gene *FBXO38* (5q32), located 839kb away from the nearest GWAS-identified risk variant, rs10793821^32^ (Table 1). For colorectal cancer, eight significant APA events were found, corresponding to eight putative susceptibility genes (Figure 2; Table S3). Of them, four were reported in previous genetic studies (Table S7). Of the remaining four genes (*FZR1, ERP29, ITCH,* and *STAT6*), only one gene, *FZR1* (19p13.3), located 2.4Mb away from the nearest index variant, rs62131228^33^, remained a significant association with colorectal cancer risk after adjusting for the index variant (Table 1). For lung cancer, we found 10 significant APA events, corresponding to seven putative susceptibility genes (Figure 2; Table S3). Of them, four were reported in previous genetic studies (Table S8). The remaining three genes not previously linked to lung cancer risk, two genes, *IVD* (5q32) and *TUBB* (6p21.33), are located more than 500Kb away from any GWAS-identified risk variants for lung cancer. Conditional analysis showed that associations with these genes remained statistically significant after adjusting for the index variants (*P* < 0.004 for all; Table 1).

**Figure 2.**
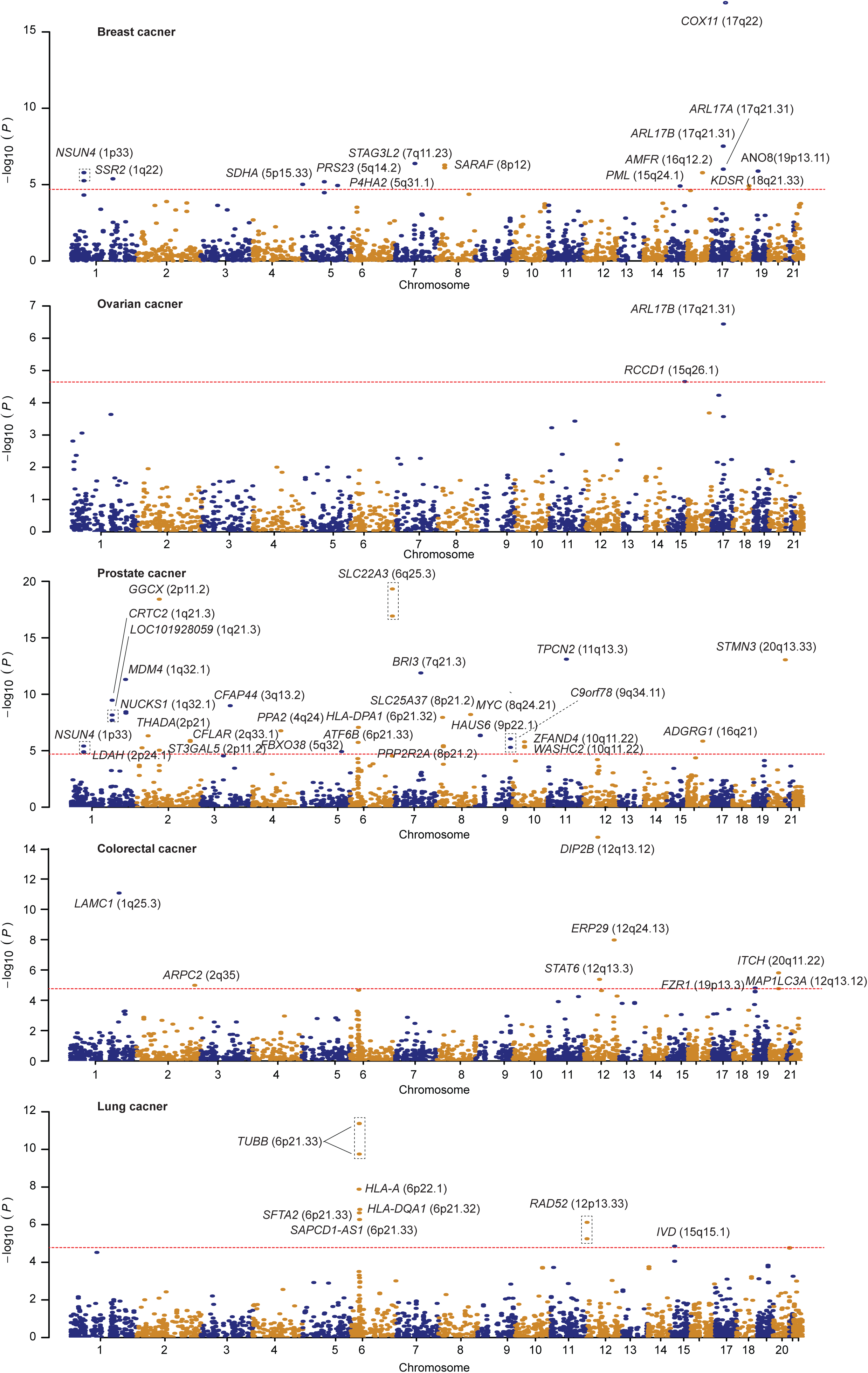
Association results from APA-WAS of breast, ovarian, prostate, colorectal, and lung cancers. Manhattan plots show association results of genetically predicted APA levels with cancer risk. Putative cancer susceptibility genes were highlighted based on the annotation of significant APA corresponding to their genes. The dashed red line denotes a threshold at Bonferroni-corrected *P* < 0.05.

**Table 1.** Putative novel susceptibility genes identified through APA-WAS for cancer of breast, ovarian, prostate, colorectum, and lung.

| Locus | Gene <sup>a</sup> | Z score | P value <sup>b</sup> | R<sup>2</sup> <sup>c</sup> | Closest index variant | Distance (Mb) <sup>d</sup> | P value after adjusting for the risk variant <sup>e</sup> |
| --- | --- | --- | --- | --- | --- | --- | --- |
| **Breast cancer** |  |  |  |  |  |  |  |
| 18q21.33 | **KDSR** | -4.38 | $1.20 \times 10^{-5}$ | 0.12 | rs17743054 | 1.8 | $2.80 \times 10^{-6}$ |
| 15q24.1 | **PML** | 4.37 | $1.23 \times 10^{-5}$ | 0.07 | rs2290203 | 1.7 | $1.23 \times 10^{-5}$ |
| 1q22 | **SSR2** | 4.60 | $4.17 \times 10^{-6}$ | 0.08 | rs4971059 | 0.83 | 0.002 |
| 8p12 | *SARAF* | -5.02 | $5.25 \times 10^{-7}$ | 0.09 | rs9693444 | 0.41 | 0.59 |
| 5p15.33 | *SDHA* | -4.43 | $9.61 \times 10^{-6}$ | 0.09 | rs62641919 | 0.009 | 0.04 |
| 19p13.11 | *ANO8* | 4.84 | $1.30 \times 10^{-6}$ | 0.09 | rs67397200 | 0.003 | 0.30 |
| **Prostate cancer** |  |  |  |  |  |  |  |
| 5q32 | **FBXO38** | -4.38 | $1.19 \times 10^{-5}$ | 0.04 | rs10793821 | 1.39 | $1.15 \times 10^{-5}$ |
| 2p11.2 | *ST3GAL5* | -4.44 | $8.99 \times 10^{-6}$ | 0.04 | rs2028900 | 0.29 | $1.03 \times 10^{-4}$ |
| 8q24.21 | *MYC* | 5.81 | $6.22 \times 10^{-9}$ | 0.02 | rs12549761 | 0.21 | $1.84 \times 10^{-10}$ |
| 10q11.22 | *WASHC2C* | 4.61 | $4.02 \times 10^{-6}$ | 0.03 | rs7075427 | 0.18 | $8.85 \times 10^{-5}$ |
| 8p21.2 | *PPP2R2A* | -4.63 | $3.60 \times 10^{-6}$ | 0.06 | rs12677206 | 0.16 | $4.43 \times 10^{-5}$ |
| 2q33.1 | *CFLAR* | 4.85 | $1.24 \times 10^{-6}$ | 0.08 | rs1861270 | 0.12 | 0.048 |
| 2p21 | *THADA* | 5.03 | $4.79 \times 10^{-7}$ | 0.06 | rs28514770 | 0.07 | $3.07 \times 10^{-5}$ |
| 16q21 | *ADGRG1* | 4.82 | $1.41 \times 10^{-6}$ | 0.05 | rs11863709 | 0.04 | $2.43 \times 10^{-6}$ |
| 20q13.33 | *STMN3* | -7.46 | $8.84 \times 10^{-14}$ | 0.04 | rs3787099 | 0.03 | $3.70 \times 10^{-17}$ |
| 1q21.3 | *LOC101928059* | -5.79 | $6.86 \times 10^{-9}$ | 0.06 | rs10127983 | 0.02 | 0.88 |
| 10q11.22 | *ZFAND4* | -4.80 | $1.61 \times 10^{-6}$ | 0.05 | rs7075427 | <0.01 | 0.16 |
| 1q21.3 | *CRTC2* | 6.28 | $3.37 \times 10^{-10}$ | 0.06 | rs10127983 | <0.01 | 0.21 |
| **Colorectal cancer** |  |  |  |  |  |  |  |
| 19p13.3 | **FZR1** | 4.32 | $1.54 \times 10^{-5}$ | 0.02 | rs62131228 | 2.4 | $5.79 \times 10^{-6}$ |
| 12q24.13 | *ERP29* | 5.72 | $1.05 \times 10^{-8}$ | 0.05 | rs3184504 | 0.58 | 0.27 |
| 20q11.22 | *ITCH* | 4.81 | $1.55 \times 10^{-6}$ | 0.02 | rs2295444 | 0.07 | 0.16 |
| 12q13.3 | *STAT6* | -4.60 | $4.15 \times 10^{-6}$ | 0.57 | rs4759277 | 0.04 | 0.04 |
| **Lung cancer** |  |  |  |  |  |  |  |
| 15q15.1 | ***IVD*** | -4.35 | $1.39 \times 10^{-5}$ | 0.02 | rs66759488 | 6.9 | $1.31 \times 10^{-4}$ |
| 6p21.33 | ***TUBB*** | 6.93 | $4.28 \times 10^{-12}$ | 0.07 | rs3094604 | 0.74 | $5.33 \times 10^{-4}$ |
| 6p21.33 | *SAPCD1-AS1* | -5.01 | $5.31 \times 10^{-7}$ | 0.02 | rs3117582 | 0.11 | 0.004 |

<sup>a</sup> Bold font denoted genes located at loci more than 500Kb away from any GWAS-identified risk variants for a cancer of interest.
<sup>b</sup> *P* value derived from APA-WAS among European-ancestry individuals; statistically significant based on a Bonferroni-corrected $P < 0.05$ for each cancer.
<sup>c</sup> Prediction performance ( $R^2$ ) was derived from APA prediction model building in normal breast, ovary, prostate, colorectal or lung tissues from the GTEx.
<sup>d</sup> Distance between a gene with the closest lead variant identified from previous GWAS in breast or ovary cancer.
<sup>e</sup> *P* value derived from association analyses in APA-WAS after adjusting for the closest lead variant of each locus.

In total, our study uncovered 58 putative susceptibility genes (total 76 significant APA events) for cancers of breast, ovary, prostate, colorectum, and lung. Of them, 25 genes were not previously reported in association with cancer risk, including seven genes at potential novel loci, which are located more than 500Kb away from any GWAS-identified risk variants (Figure 3A; Table 1). For pancreatic cancer, none of the associations remained statistically significant adjusting for multiple comparisons. In addition to our reported genes using the stringent threshold of Bonferroni-correction, we also identified many genetically predicted APA associated with cancer risk at the nominal *P* < 0.05 (Table S3). To further assess the likelihood of shared causal variants between APA-WAS and GWAS, we performed colocalization analyses on all 76 significant APAs identified in our APA-WAS using COLOC ^34^ (see **Methods**). Among them, 17 unique genes (from 20 APAs, 26.3%) showed evidence of co-localization (Table S9). This finding overall aligns with previous observations from the colocalization analysis of trait-associated GWAS and 3’aQTL signals^21^.

**Figure 3.**
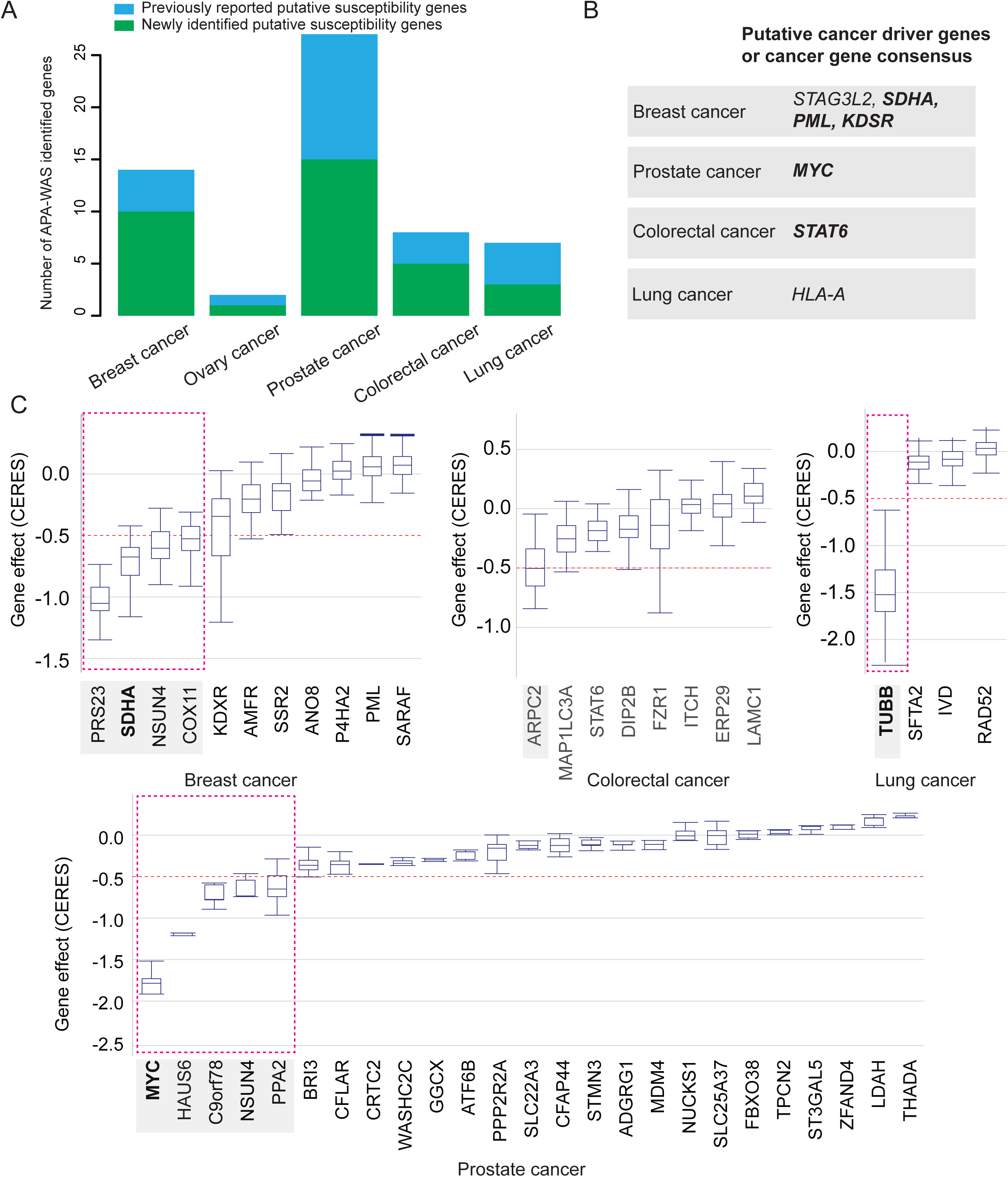
Functional evidence of oncogenic roles for the putative susceptibility genes identified from APA-WAS of breast, ovarian, prostate, colorectal, and lung cancer. A) A barplot shows the number of target genes identified for breast, ovary, prostate, and lung cancers. The number of previously unreported genes and reported genes are denoted by green and blue, respectively. B) A list of putative cancer driver genes or cancer gene consensus for previously unreported genes (bold font) and reported genes C) Boxplots show effects of APA-WAS identified genes on cell proliferation using experimental data from CRISPR screens (see Methods). Dashed red boxes highlight a total of 10 genes, including three previously unreported genes (bold font), which showed evidence of essentiality on cell proliferation based on a cutoff of median CERES values < −0.5.

Of note, we found that eight putative susceptibility genes at five loci were commonly implicated in cancers of breast and ovary (*ARL17B* at 17q21.31), breast and prostate (*NSUN4* at 1p33), prostate and colorectal (*STMN3* at 20q13.33), prostate and lung (*HLA-DPA1* at 6p21.32; *HLA-DQB1* at 6p21.32; *HLA-DQA1* at 6p21.33) and colorectal and lung (*HLA-DRB6* at 6p21.32; *HLA-A* at 6p21.33) (see **Methods**; Table S3-S8). These results provide evidence of these putative susceptibility genes underlying cancer pleiotropy for shared cancer risk.

In addition, we conducted joint TWAS analysis based on multiple tissues models using two approaches, multivariate adaptive shrinkage (MASH)^35^ and the aggregated Cauchy association test (ACAT)^36^ (see **Methods**). Pooling the results across the six cancer types, we have detected total 2,385 and 121 significant associations using MASH and ACAT, respectively. Of our identified genes, 46 (79.3% of 58) were also detected by both MASH and ACAT approaches (Table S10). It’s worth noting that the majority of associations detected by the ACAT method were encompassed within the results of the MASH method. The substantial increase in the number of associations identified by MASH can be attributed to its utilization of a comparatively less stringent statistical cutoff, known as the local false sign rate (LFSR). Nonetheless, additional research is imperative to validate the findings.

### Functional evidence of oncogenic roles for the APA-WAS-identified genes

Functional enrichment analyses using the Ingenuity Pathway Analysis tool (IPA) showed that these 58 newly identified putative susceptibility genes were significantly enriched in cancer function category and 50 of them have been implicated in carcinogenesis (*P* < 0.05; Table S11). We further examined whether they were implicated in carcinogenesis from previous studies, such as cancer driver genes^37, 38^, or Cancer Gene Census (CGC)^39^ (see **Methods**). We found evidence for seven genes as potential cancer drivers and/or CGC, including *SDHA, PML, KDSR, and STAG3L2* for breast cancer, *MYC* for prostate cancer, *STAT6* for colorectal cancer, and *HLA-A* for lung cancer (Figure 3B; Table S11).

We further explored potential functional roles of putative susceptibility genes identified in our study using CRISPR screen silencing data to investigate gene essentiality on cell proliferation in breast (n=40), ovarian (n=47), prostate (n=5), and lung (n=116) cancer relevant cell lines (see **Methods)**. Using a cutoff of median CERES Score < −0.5 ^40, 41^, we found evidence of essentiality for four genes (*RPS23, SDHA, NSN4, and COX11*) for breast cancer cell proliferation; five genes (*MYC*, *HAUS6*, *C9orf78*, *NSUN4*, and *PPA2*) for prostate cancer cell proliferation; *ARPC2* for colorectal cell proliferation, and *TUBB* for lung cell proliferation (Figure 3C; Table S11).

### Putative susceptibility genes supported by 3’aQTL analysis and functional genomic data

To verify susceptibility genes identified by APA-WAS analyses, we additionally evaluated 3’aQTL results from the lead variant in each of the predicting models (see **Methods**). We found that most of them (97.4%) were associated with these genes at nominal *P* < 0.05, providing additional support for our discovery (Table S12). Specifically, 3’aQTL analysis showed significant association for those unreported genes with the above functional evidence of oncogenic roles: *SDHA* (lead variant rs113742171), *PML* (rs71137385), and *KDSR* (rs6567326) in breast cancer*, MYC (*rs16902085) in prostate cancer, *STAT6* (rs324015) in colorectal cancer, and *TUBB* (rs9262120) in lung cancer (Figure 4).

**Figure 4.**
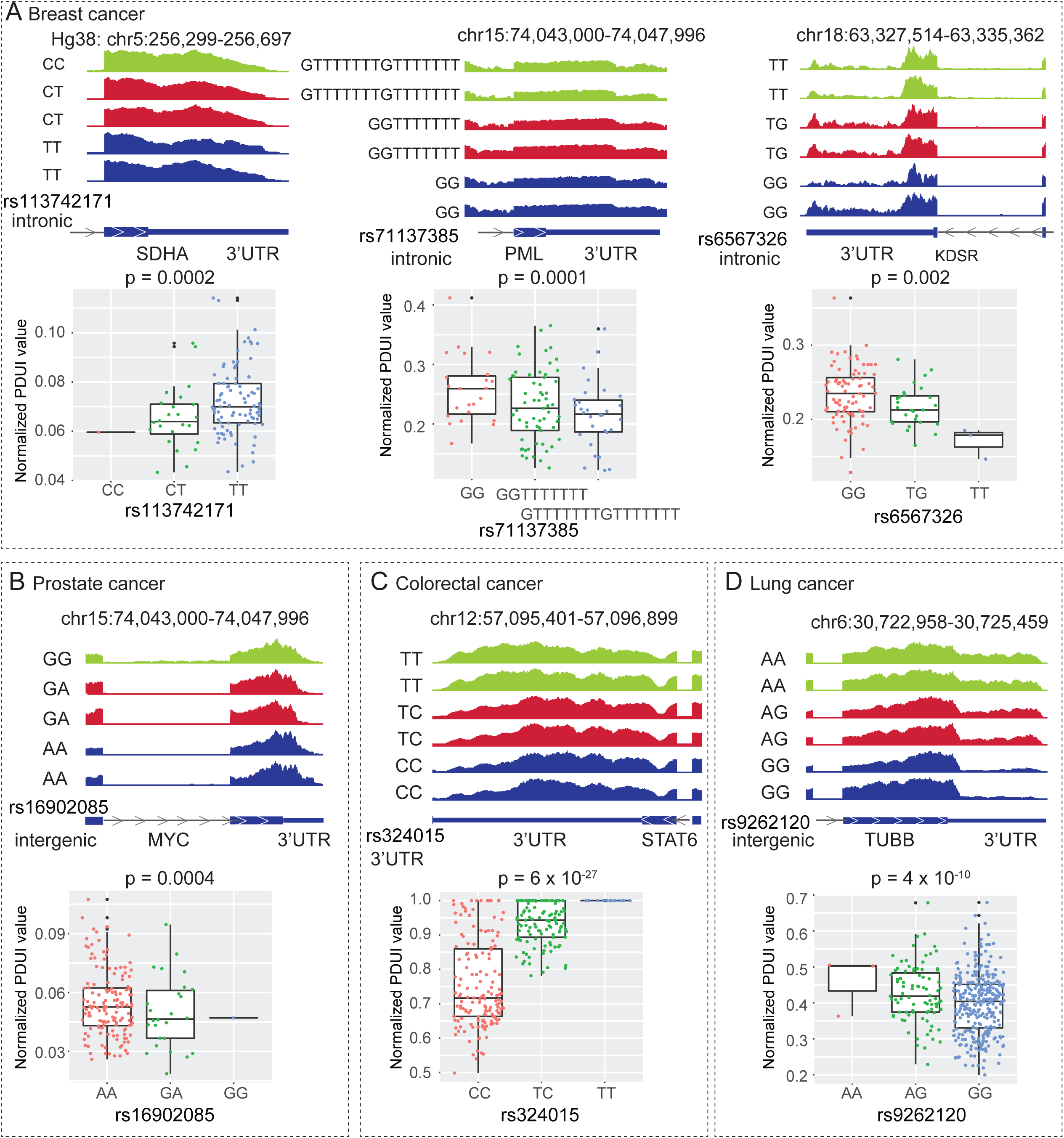
Putative susceptibility genes supported by 3a’QTL analyses. The highlighted unreported genes with functional evidence of oncogenic roles for A) breast cancer, B) prostate cancer, C) colorectal cancer and D) lung cancer, are supported by 3a’QTL analysis for the lead variants included in prediction models of APA-WAS identified genes. The top panels illustrate RNA-seq coverage track for the 3’UTR of a gene of interest from selected samples with different genotypes, which serve as an illustrative means to convey the overarching pattern for the specified PDUI value. The gene structure, including 3’UTR, was annotated based on the Refseq database. Boxplots indicate the normalized PDUI values for samples within three groups defined by genotypes.

To further search evidence of regulatory mechanisms underlying the identified genes, we performed extensive functional annotation analysis to characterize candidate functional variants in strong LD with the lead variants in the prediction model (see **Methods**). We evaluated the functionalities of total 1,430 variants in strong LD (R^2^ > 0.8) with the 76 lead SNP in the prediction model for each gene. Of them, we found a total of 33 genes (56.9% of 58 genes) that were the closest genes for these putative regulatory 3’aQTL variants (Table S13). In particular, 18 genes were associated with variants that are located in their 3’UTR. By comparison, only one gene, *NSUN4*, was associated with variants that are located in its 5’ UTR, strongly indicating that 3’aQTL variants for our identified closet genes were significantly enriched in their 3’ UTR (Binomial test, *P* < 2.2 × 10^-16^). These results provide strong evidence for many of our identified genes supported by 3’aQTL analysis and functional genomics data.

### Luciferase Reporter Assays for putative 3’UTR functional variants and their target genes

To investigate potential mechanisms of the lead variants in regulating their target genes, we selected five APA-WAS identified genes and evaluate if these genes may be regulated by their corresponding putative 3’UTR functional variants, including rs324015 (*STAT6*), rs2280503 (*DIP2B*), rs1128450 (*FBXO38*), rs145220637 (*LDAH*) and rs72550303 (*AMFR*) (Table S13, Figure 5A, see **Methods**). We performed luciferase reporter assays for these variants in 293T and target cancer cell lines selected based on the genes identified in the analysis of that particular cancer (CRC: *STAT6* and *DIP2B* in HCT116; and prostate cancer: *FBXO38* and *LDAH* in VCaP; and breast cancer: *AMFR* in MDA-MB-231). Our results show that, compared to the reference alleles, fragments containing risk alleles significantly increased the luciferase signals of *STAT6, DIP2B*, *FBXO38* and *LDAH* in both 293T and target cancer cell lines (Figure 5B). These results are in line with the APA-WAS and 3’aQTL findings (Figure 5B). However, when investigating the activities of *AMFR*, we failed to detect a consistent pattern between alternative and reference alleles in both cell lines.

**Figure 5.**
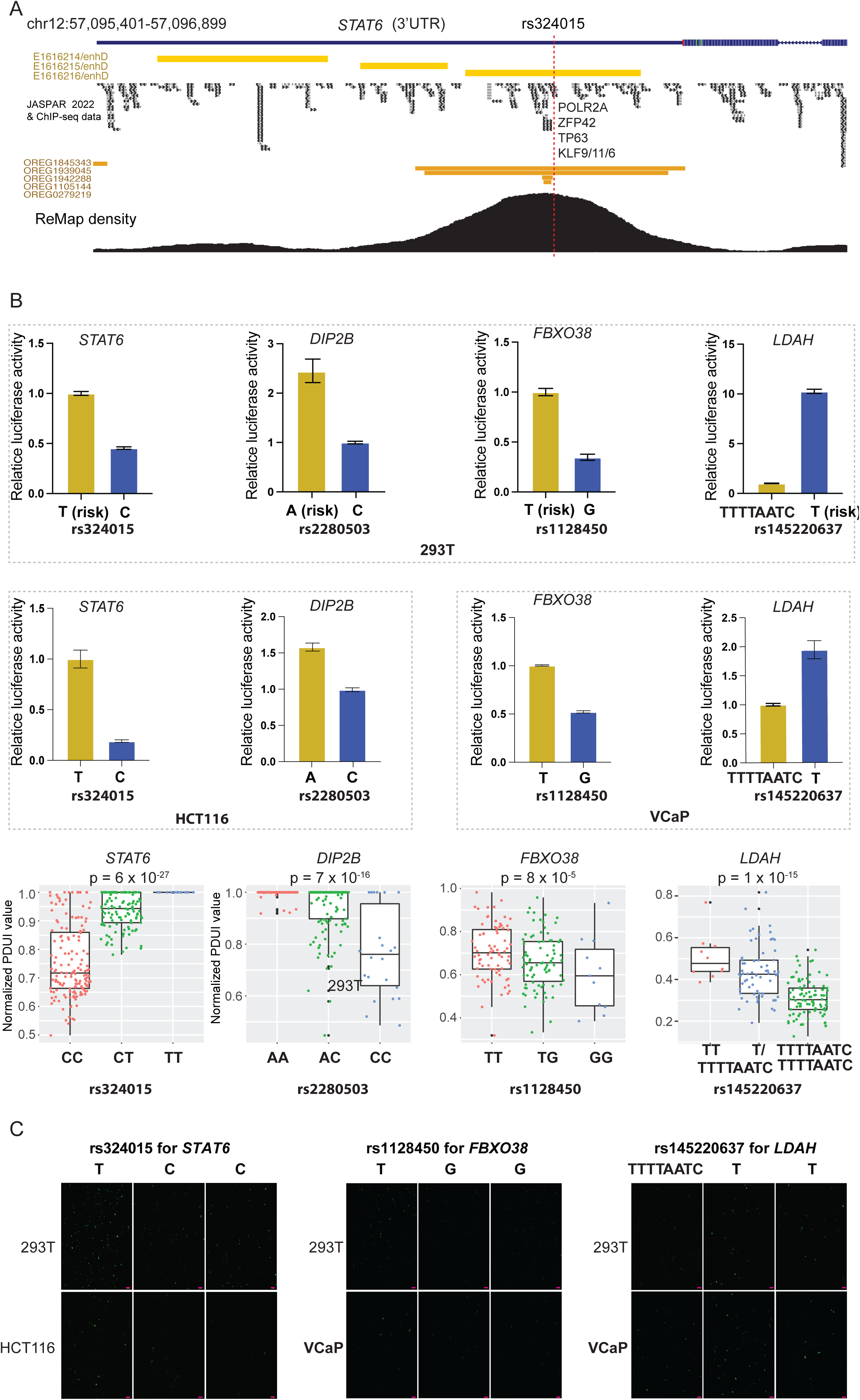
Alternative alleles affecting target genes’ activity using Dual-Luciferase reporter and EGFP assays. A) Functional genomics analysis showed that the *STAT6* gene potentially regulated by the putative regulatory 3’ UTR variant, rs324015 (i.e., enhance, transcription factor motif and ChIP-seq binding site, ReMap). B) Luciferase assays were conducted to investigate 3’UTR variants affecting post-transcriptional activities in 293T cell line (top panel) and target cancer cell lines (bottom panel). Barplot showed that alternative alleles of the 3’UTR variants, rs324015 (*STAT6*), rs1128450 (*FBXO38*), rs2280503 (*DIP2B*) and rs145220637 (*LDAH*), could significantly change their luciferase activities compared to reference alleles in both 293T and target cancer cell lines. The error bar represents the mean and standard deviation (SD) of the promoter activity of a target gene of interest. C) The expression of EGFP of WT and mutant plasmid for *STAT6* (risk allele: T) transfected cells was observed under fluorescence microscopy in 293T and HCT116 cells; the expression of EGFP of WT and mutant plasmid for *FBXO38* (risk allele: T) transfected cells was observed under fluorescence microscopy in 293T and Vcap cells; and the expression of EGFP of WT and mutant plasmid for *LDAH* (risk allele: T) transfected cells was observed under fluorescence microscopy in 293T and Vcap cells. Mutant plasmids for transfected cells were all presented with two fluorescence images Scale bar: 50 μm.

Moreover, we conducted an exploration into the lead variants, namely rs324015, rs1128450, and rs145220637, using an EGFP fluorescence assay in both 293T and the target cancer cell lines (except for DIP2B, which was excluded due to its longer 3’UTR). We transfected *STAT6* WT and mutant reporter plasmids into both 293T and HCT116 cell lines. Similarly, *FBXO38* and *LDAH* WT and mutant reporter plasmids were transfected into 293T and Vcap cell lines (see **Methods**). We showed that risk alleles of rs324015, rs1128450 and rs145220637 significantly increased fluorescence intensities *STAT6*, *FBXO38* an*d LDAH,* compared to the reference alleles in in both 293T and target cancer cell lines. These findings further strengthen the evidence that the target gene of interest is under the regulation of its 3’UTR variants, rs324015, rs1128450 and rs145220637 (Figure 5C).

### Functional assays for putative susceptibility genes

We next conducted small interfering RNA (siRNA) transfection experiments and *in vitro* functional assays for *STAT6* in HCT116 cells and *FBXO38* and *LDAH* in DU145 cells (see **Methods**). Notably, for the purpose of cell scratching assays, we encountered limitations in utilizing Vcap cells, which subsequently led us to opt for the prostate cancer cell line DU145 for our subsequent cellular assessments. The effectiveness of their silencing was confirmed through qPCR assays, which demonstrated substantially lower expression levels of these genes in cells transfected with siRNAs in comparison to cells in the si-NC group (Figure 6A; *P* < 0.01 for all cases). Furthermore, the results from cell proliferation CCK8 assays unveiled significant inhibitory effects on cell proliferation in both DU145 and HCT116 cells upon the silencing of *FBXO38, LDHA*, and *STAT6* genes (Figure 6B). Consistently, results from the transwell assay revealed a substantial reduction in both cell migration and invasion post-silencing of these three genes compared to the control group (Figure 6C). Notably, among the genes examined, *FBXO38* exhibited the most pronounced impact on cell migration and invasion in DU145 cells. Furthermore, wound healing efficiency was noticeably diminished in comparison to control cells, confirming the capacity of these genes to influence cell migration (Figure 6D).

**Figure 6.**
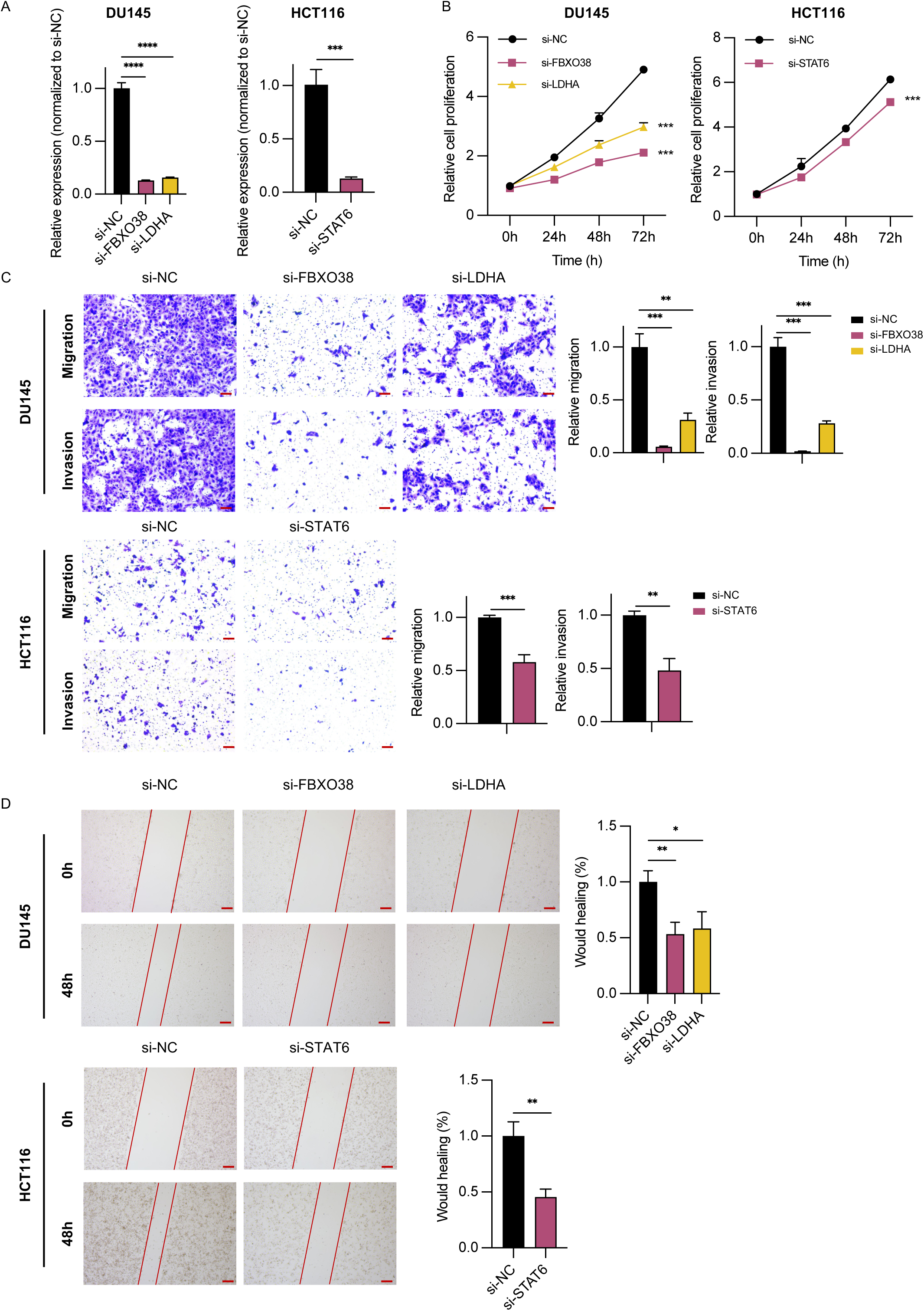
*In vitro* Functional assays of *FBXO38, LDHA*, and *STAT6*. A) RT-qPCR results of relative expression of *FBXO38* and *LDHA* were knockdown in DU145 cells, and STAT6 was silenced in HCT116 cells. B) The proliferation ability of DU145 and HCT116 transfected with si-FBXO38, si-LDHA, si-STAT6 and si-NC was detected by CCK8 proliferation assay. C) The results of transwell experiments revealed the migration and invasion abilities of DU145 and HCT116 were inhibited after transfection. Statistical results are quantification of cells across the membrane. Images were taken for each membrane. Scale bar= 20 µm. D) Wound healing assay of cell migration and invasion abilities of gene silenced cells. Assays were performed in DU145 cells transfected with si-NC, si-FBXO38 and si-LDHA, and in HCT116 cells transfected with si-NC and si-STAT6. Representative images (magnification × 4) are shown after 0 and 48 h. Unhealed areas were measured and quantified by ImageJ software. Scale bar= 100 µm. Three independent experiments were performed. Data are mean ± SEM; *: *P* < 0.05, **: *P* < 0.01, ***: *P* < 0.001, ****: P < 0.0001.

## Discussion

To expand the scope of traditional TWAS, this is the first study to systematically evaluate associations of genetically predicted APA levels with risk of breast, ovarian, prostate, colorectal, lung, and pancreatic cancers. We identified 25 genes not previously linked to cancer risk, including seven genes identified in regions not located in any of the risk loci revealed previously by GWAS. Our findings highlighted the potential of APA-WAS in uncovering novel associations and highlights genetically regulated the 3’ UTR APA in contributing susceptibilities in human common cancers. Furthermore, through analysis of multiple cancers, our results revealed that eight putative susceptibility genes at five loci were commonly implicated in different cancers. These results provide further evidence of these putative susceptibility genes underlying cancer pleiotropy for shared cancer risk.

There is strong evidence in support of significant roles for multiple putative susceptibility genes uncovered in our study in the etiology of cancer. For example, six have strong evidence of oncogenic roles: *SDHA, PML, and KDSR* (breast cancer)*; MYC* (prostate cancer); *STAT6* (colorectal cancer) and *TUBB* (lung cancer). These genes, except for *TUBB,* have been observed to be frequently mutated in cancer tissues, annotated as potential cancer driver genes^37, 38, 39^. Other studies provided additional evidence of their oncogenic functions, including *SDHA* associated with younger age at diagnosis and low-grade histology in breast cancer^42^ and *PML* playing a regulatory role in the TGF-beta signaling pathway^43, 44, 45^, *KDSR* involved cellular metabolism^46^; *MYC* driving tumorigenesis in prostate cancer^47, 48, 49, 50, 51^; *STAT6* promoting intestinal inflammation and tumorigenesis^52^; and *TUBB* correlated with prognosis among lung adenocarcinoma patients^53^. In particular, In particular, our functional assay has unveiled that the depletion of *STAT6, DIP2B*, and *LDAH* yields a noteworthy capability to significantly suppress cell proliferation efficiency. Furthermore, it substantially diminishes the effectiveness of cell migration and invasion in both HCT116 and DU145 cell lines. These findings offer robust evidence of the pivotal roles played by these genes in CRC etiology and biology.

Both TWAS and APA-WAS illuminate distinct facets of the intricate interplay between genetics and gene regulation. While TWAS predominantly unravels the intricate connections between genetic variants and gene expression levels, APA-WAS investigations cast a spotlight on how these variants influence APA levels, consequently impacting mRNA stability, isoforms, and even potentially steering protein products. In our study, at a Bonferroni-corrected *P* < 0.05, we observed that approximately 22% of the identified APAs exhibit a significant correlation with mRNA expression and 25% of them have significant eQTLs (Table S12). Intriguingly, only 53 % of these correlated APAs have relaxed eQTL signals at a nominal *P* < 0.05. This observation overall aligns with prior research, which has underscored that, within the same genes, 3’aQTLs typically manifest as a significant proportion independent from eQTLs^21^. These findings underscore the pivotal role of APA-WAS as an additional avenue for uncovering susceptibility genes that might elude detection via conventional TWAS approaches.

Importantly, the tissue-specific nature intrinsic to APA, as compared to gene expression, signifies that APA events harbor heightened sensitivity in the identification of susceptibility genes, bolstering accuracy. Through the amalgamation of insights garnered from both methodologies, a more encompassing understanding of the intricate molecular mechanisms that underpin genetic associations with traits and diseases emerges. This synergistic approach facilitates a more holistic comprehension of gene regulation, advancing our knowledge within the realms of cancer contexts.

We have demonstrated robust predictive performance for our identified putative susceptibility genes, with median R^2^ values of 0.12, 0.07, 0.04, 0.06, and 0.24 for breast, prostate, colon, lung, and ovarian cancers, respectively (Table S3). Significantly, a majority of the genes we identified found support through 3’aQTL analysis of the lead variants within the expression prediction model. Furthermore, we observed that over half of these genes were in close proximity to putative regulatory 3’aQTL variants, particularly those situated in the 3’ UTR regions. Through luciferase reporter assays targeting putative 3’ UTR functional variants, we effectively demonstrated that the risk alleles of these variants exerted significant alterations in the post-transcriptional activities of their respective target genes, compared to the reference alleles. While the findings offer compelling evidence of 3’UTR functional variants, a more comprehensive investigation is warranted. This could involve employing CRISPR-Cas9 mediated knockout DNA fragments, incorporating the alleles of the variants of interest, to assess their effects on APA site usage. It’s worth highlighting that these variants may not necessarily be located within differentially transcribed 3’ UTR regions, such as the case of rs324015 for *STAT6*, as their association with APA is established via our 3’aQTL analysis. This suggests a potential dual role for them in the regulation of both APA and target gene activity. However, it’s important to acknowledge that both 3’aQTL and APA-WAS provide statistical evidence of these genetic variants linking to genes via correlations with APA levels. Nevertheless, characterizing regulatory 3’aQTL variants remains challenging, especially considering that a considerable number of them reside in intronic or other cis-regulatory elements (Table S13). It’s plausible that gene regulation may extend to distal mechanisms involving functional variants interacting with 3’ UTR enhancers. This underscores the need for additional epigenetic data, coupled with downstream experiments, to further unravel the underlying mechanisms driving these observed associations. Furthermore, APA plays a pivotal role in intricate post-transcriptional gene regulation, encompassing alterations in 3’ UTR length, poly(A) motifs, RNA secondary structures, and RNA-binding protein (RBP) binding sites. These modifications culminate in effects on competitive endogenous RNA (ceRNA) dynamics by sequestering miRNAs, thereby influencing mRNA stability, translation efficiency, and protein subcellular localization^21, 54, 55, 56^. To comprehensively advance our understanding of disease susceptibility mechanisms, it is imperative to delve into the intricate interplay of genetic-based 3’ UTR and APA regulations within complex biological processes.

In conclusion, we conducted the first APA-WAS for six major cancers and identified a large number of putative susceptibility genes not previously linked to cancer risk. Our study highlights genetically regulated the 3’ UTR APA in contributing cancer risk and provides additional insight into the genetic susceptibility of these common cancers.

## Methods

### Data resources

We used whole genome sequencing (WGS) in blood samples and RNA sequencing (RNA-seq) data generated in normal tissues from the GTEx project (version 8) as the referenced data in our model building of genetically predicted APA levels^57^.

WGS data were sequenced using the Illumina HiSeq X platform (except the first batch by Illumina HiSeq 2000) by the Broad Institute’s Genomics Platform on DNA samples from 838 donors to a median coverage of ∼32X. RNA samples from 54 tissue sites were sequenced using Illumina TrueSeq to generate transcriptome profiling data. All raw RNA-Seq files (BAM) and genotype files (VCF), along with sample attributes and subject phenotype information, were downloaded from the dbGap and the Google cloud (Accession No. phs000424.v8.p2). We downloaded BAM files with mapped RNA-seq data for all 49 tissues with at least 50 samples from European individuals who also had whole genome sequencing data available in GTEx (n = 11,722).

Summary statistics of GWAS data of European descendants for breast, ovarian, prostate, and lung cancers have been released from their consortia (Table S1). GWAS data for breast cancer were downloaded from the Breast Cancer Association Consortium (BCAC)^24^. BCAC is an international, multidisciplinary consortium designed to identify genetic susceptibility factors that are related to the risk of breast cancer. BCAC has generated GWAS data for a total of 133,384 cases and 113,789 controls from European descendants. For prostate cancer, GWAS data of 79,194 cases and 61,112 controls from European descendants were released from the Prostate Cancer Association Group to Investigate Cancer Associated Alterations in the Genome (PRACTICAL)^32^. For colorectal cancer, the Genetics and Epidemiology of Colorectal Cancer Consortium (GECCO), the Colorectal Cancer Transdisciplinary Study (CORECT) and the Colon Cancer Family Registry (CCFR) generated GWAS data of 58,131 cases and 67,347 controls^9^. For ovarian cancer, GWAS data were downloaded from the Ovary Cancer Association Consortium (OCAC)^3^. GWAS data were generated from 63 genotyping project/case-control sets with 22,406 invasive epithelial ovary cancer (EOC) cases and 40,941 controls from European descendants. For lung cancer, GWAS data were downloaded from the Transdisciplinary Research of Cancer in Lung of the International Lung Cancer Consortium (TRICL-ILCCO) and the Lung Cancer Cohort Consortium (LC3) totaling 29,266 cases and 56,450 controls from European descendants^4^. For pancreatic cancer, GWAS data were downloaded from the Pancreatic Cancer Cohort Consortium (PanScan) and the Pancreatic Cancer Case-Control Consortium (PanC4) totaling 8,275 cases and 6,723 controls from European descendants^58^.

We compiled lists of cancer driver genes from two previous studies^59, 60^ and CGC^39^ from the COSMIC website. To investigate the effect of an individual gene on essentiality for proliferation and survival of cancer cells, the dataset “CRISPR DepMap 21Q3 Public measured by CERES” was downloaded from the DepMap portal. To characterize putative cancer susceptibility genes, *cis-*eQTL association data of each tissue for GWAS-identified risk variants in these major cancer types were downloaded from GTEx Google Cloud resources.

### Genotype data processing

Genotype data of European descendants (n=660) were extracted from the VCF file in the WGS data. Genetic variants with minor allele frequency (MAF) < 5%, Hardy-Weinberg equilibrium (HWE) *P* value < 10^-4^, and missing data > 5% were excluded. Multi-allelic SNPs and SNPs with alleles A/T, T/A, C/G, and G/C were also excluded. After the above multiple quantity control steps, for each cancer type, only SNPs with summary statistics data available were remained. Finally, included for prediction model building were 5,610,175 SNPs for breast tissue, 5,631,452 SNPs for ovarian tissue, 5,673,856 SNPs for prostate tissue, 4,602,635 SNPs for colon transverse tissue, 4,556,226 SNPs for lung tissue and 3,809,159 SNPs for pancreases tissue, respectively.

### Model building of genetically predicted APA levels in normal breast, ovarian, prostate, colon transverse, pancreas, and lung tissues

We used BAM files with mapped RNA-seq data generated in normal tissues of breast, ovary, colon transverse, prostate, and lung to quantify APA level by using percentage of PDUI estimated from DaPars v2.0^21^. For each tissue, an APA event was removed if it was more than 5% missing values among all subjects. We performed quantile normalization to transform the quantified PDUI values of APA for each sample to the same distribution. We estimated probabilistic estimation of expression residuals (PEER) factors using the normalized APAs to correct for batch effects and experimental confounders in our downstream prediction model building^61^. For each tissue type, the number of PEER factors was determined as a function of sample size (N): 15 factors for N<150, 30 factors for 150≤N<250, 45 factors for 250≤N<350, and 60 factors for N≥350, as previously suggested^61, 62^.

We built genetically predicted models for each APA of each tissue based on the processed genotype data and measured APA levels. We first used linear models to generate residuals of APA levels (normalized PDUI values) by adjusting age, sex, RNA integrity number (RIN), PEER factors, and top three genotype principal components. Inverse quantile normalization was conducted for the generated residuals. We further used all genetic variants flanking ±1Mb region to train the elastic-net model^11^. Finally, the prediction performance of each APA prediction model satisfied with R^2^ > 0.01 (10% correlation) at *P* < 0.05, were used for downstream association analysis.

### Association analysis between genetically predicted APA levels and cancer risk

We analyzed the association between genetically predicated APA levels and cancer risk using the method from the S-PrediXcan tool (see the formula below).

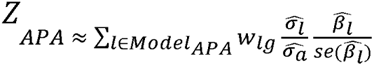

In this formula, *w_lg_* is the weight of a genetic variant *l* for predicting the levels of APA *g*. 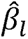 and 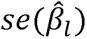 are the association regression coefficient and its standard error for a genetic variant *l* in GWAS, and 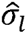 and 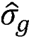 are the estimated variances of a variant *l* and the predicted levels of APA *g*, respectively. A Bonferroni-corrected *P* < 0.05 was used for identifying significant APAs associated with cancer risk (adjusting for 0.05/2,556 tests for breast cancer; 0.05/2,363 for ovarian cancer; 0.05/2,631 for prostate cancer; 0.05/2,913 for colorectal cancer; 0.05/3,098 for lung cancer and 0.05/1,736 for pancreatic cancer).

We additionally conducted conditional analyses by adjusting for the nearest GWAS risk signal (the lead SNP, with the strongest association with cancer risk in the locus). For each variant included in the model of genetically predicted APA levels, GCTA-COJO analyses^63^ was performed to calculate the statistical significance with cancer risk after adjusting for the nearest lead variant. We further conducted S-PrediXcan analyses based on the adjusted statistics to investigate the genetically predicted APA levels in association with cancer risk.

### Colocalization analysis between 3’a QTL and GWAS signals

COLOC analyses were conducted to evaluate the likelihood of shared causal variants between 3’aQTL and GWAS signals for APA/gene regions identified by our APA-WAS^34^. For each genetic variant located within the defined region of the identified APA (spanning +/-500Kb), we examined summary statistics from 3’aQTL results originating from GTEx and GWAS association statistics for the relevant cancer types of interest. To calculate the posterior probability of colocalization, we employed default priors and utilized the coloc.abf function. The coloc tool provides posterior probabilities (PP) for five hypotheses, with our primary focus placed on PP.H4. This particular posterior probability (PP.H4) corresponds to hypothesis H4, estimating the likelihood of a singular causal variant being associated with both protein levels and the traits of interest. Our criterion for determining a gene to potentially host a shared causal variant from both 3’aQTL and GWAS was based on the coloc PP.H4 value > 0.5.

### Joint TWAS analysis based on multiple tissue models

We partitioned the samples based on sex to facilitate the use of sex-specific cross-tissue analyses, specifically for breast, ovary, and prostate tissues. Within each tissue and sex subset, we imposed a prerequisite of including a minimum of 50 samples to ensure robustness in subsequent analyses (32 tissues for European females and 45 for European males). To quantify the APA levels, we utilized the percentage of Percent Distal UTR Inclusion (PDUI), as estimated through DaPars v2.0. The additional samples underwent the same quality control and preprocessing procedures as those employed for the six core tissues in our original analysis. Following this, we built distinct single-tissue models for these additional tissues, following the identical methodology delineated in our initial single tissue analysis. We then evaluated the association between genetically predicated APA levels and cancer risk of interest using the S-PrediXcan approach described in the preceding section.

Furthermore, we used two methods to joint TWAS combining information across multiple tissues, MASH and ACAT. MASH facilitates the estimation of effects for each APA-WAS within each tissue, accommodating both the sharing of effects across tissues and the tissue-specific aspects of APA-WAS effects. We utilized the Z-score table generated from all single-tissue APA-WAS models pertaining to the focused tissues, and conducted MASH analysis using the ‘mashr’ R package. Under the MASH method, APAs possessing a local false sign rate (LFSR) below 0.05 divided total tested APAs were classified as significant APAs. The ACAT method combines *P* values from single-tissue TWAS results across all tissues. APAs with a Bonferroni-corrected *P* value < 0.05 were considered as significant APAs using the ACAT method.

### Pathway enrichment analysis for the identified putative cancer susceptibility genes

IPA tool was used to evaluate the functional enrichment in the gene function category and biological pathways for the identified putative cancer susceptibility genes. We also examined if a gene was annotated as cancer-related function based on its involved cancer molecules from the top list on Diseases and Disorders category.

### Effect of gene silencing on cell proliferation

For each gene of interest, we calculated a median CERES value (measuring the essentiality of gene silencing on cell proliferation) of the relevant cells for each cancer type: breast, ovary, colorectal, prostate, and lung for each gene, based on the gene-dependency levels from CRISPR-Cas9 essentiality data from the DepMap portal. We used a cutoff CERES value = −0.5 as the evidence of essentiality for an investigated genes following previous literature^40^.

### Characterization of 3’aQTL analysis

We examined associations between APA-WAS-identified genes and the lead variant (i.e., the variant showing the strongest association with cancer risk) in each predicted model of these significant genes based on previous 3’aQTL analysis using data in GTEx^64^. We analyzed and characterized 3’aQTL results for each cancer of breast, colon transverse, lung, ovary, pancreas, and prostate^64^. Associations from the 3’aQTL analysis for these lead variants were extracted and summarized in Table S12.

### Functional annotation of genetic variants

Functional annotation for the lead variants in prediction models of APA-WAS-identified genes was analyzed based on HaploReg v4 database^65^ and the UCSC Genome Browser. We used data from European populations from the 1000 Genomes project to identify genetic variants in strong LD (R^2^ > 0.8) for these lead variants. We further conducted the functional annotation of each variants including genomic location, chromatin states from chromHMM annotation, DNase I hypersensitive sites and TF ChIP-seq binding peaks, using data from both ENCODE and Roadmap projects and RegulomeDB^66^.

### Cell lines and cell culture

HCT116 and DU145 cell lines were purchased from the American Type Culture Collection and cultured in RPMI 1640 and DMEM medium (Gibco) respectively. VCaP, 293T and MDA-MB-231 cell lines were purchased from the National Collection of Authenticated Cell Cultures (China) and maintained in DMEM medium. The detailed cell culture procedure was carried out as detailed previously^67^.

### Plasmids construction, Dual-luciferase reporter assay and EGFP fluorescence detection

We focused on putative 3’UTR regulatory variants for experimental investigation based on functional annotation of genetic variants in strong LD (R^2^ > 0.8) for these lead variants (see the above functional annotation section). The five 3’UTR variants, rs324015 (*STAT6*), rs1128450 (*FBXO38*), rs2280503 (*DIP2B*) and rs145220637 (*LDAH*) and rs72550303 (*AMFR*), were selected based on the uniqueness of a 3’UTR variant associated with a target gene with additional support from functional evidence (Figure 5A, Table S13). The target gene expression efficiency of functional variants was evaluated using luciferase or enhanced green fluorescent protein (EGFP) reporter assays. The 3’ UTR sequences of *STAT6, AMFR, FBXO38, DIP2B*, and *LDAH* genes containing their 3’UTR variants were cloned into PGL3-promoter vector (Promega). The alternative allele of the individual variant in the 3’ UTR region for each gene construction was introduced into the plasmid using site-directed mutagenesis. The 3’ UTR sequences of *STAT6, FBXO38*, and *LDAH*, along with their corresponding mutant sequences, were cloned separately into PEGFP-C1 vectors (Clontech). These plasmids were meticulously constructed by RealGene Bio-tech (Shanghai) and subsequently validated through sequencing.

For Dual-luciferase reporter assay, 293T, HCT116, VCaP and MDA-MB-231 cells were seeded into 24-well plates, and each constructed luciferase reporter plasmid and pRL-TK transfection control plasmid were transfected into the cells using the Lipofectamine 2000 reagent (Invitrogen). *STAT6* and *DIP2B* luciferase reporter and mutant plasmids were transfected into 293T and HCT116 cells. *FBXO38* and *LDAH* luciferase reporter and mutant plasmids were transfected into 293T and VCaP cells. AMFR luciferase reporter and mutant plasmids were transfected into 293T and MDA-MB-231 cells. After 36h incubation, the cells were collected for Dual-luciferase reporter assay as we previously reported^7, 12^. All reporter assays were performed in triplicate and repeated in two-three independent experiments.

For EGFP assay, the *STAT6* EGFP reporter and its corresponding mutant plasmids were introduced into 293T and HCT116 cells. Similarly, the *FBXO38* and LDHA EGFP reporter and mutant plasmids were transfected into 293T and Vcap cells. After 48hr, the cells were seeded into confocal dish. When the cells attached, EGFP fluorescence was examined under laser confocal microscope (Leica sp8). Fluorescence detection was performed as described previously^68^.

### Cell siRNA transfection and knockdown efficiency detection

The day before transfection, the cells were inoculated in 6-well plates. When the confluence of the cells reached 50%, the operation was performed according to the instructions of the Lipofectamine^TM^ 3000 transfection reagent (Invitrogen). Negative control (NC), si-FBXO38 and si-LDHA were transfected in the DU145 cell line, and NC, si-STAT6 were transfected in the HCT116 cell line, respectively. The siRNAs were purchased from Genepharma. After 48h transfection, the expression levels in the transfected cells were detected by RT-qPCR. Total RNA was extracted from prostate and colon cancer cell lines using AG RNAex Pro Reagent (AGbio) and reverse transcribed into cDNA. Primers used are listed bellowed: GAPDH_F: ACCACAGTCCATGCCATCAC, GAPDH_F: TCCACCACCCTGTTGCTGT; STAT6_F: AGCCCAAGGATGAGGCTTTC, STAT6_R: AATCAGGGGCCATTCCAAGG; FBXO38_F: TGCGAGTTGTGAGAGTTGTAGA, FBXO38_R: GGCCATATAGCTGTTCAACAT; LDAH_F: CTCCCGGTAATTCGTGCCTT, LDAH_R: CAGCACAAAAGTGGAGTGGC. The relative expression levels of *STAT6*, *FBXO38*, and *LDHA* were calculated by the 2^-ΔΔCt^ method using GAPDH as an endogenous control.

### Cell viability assay

After transfection, the DU145 and HCT116 cells were collected, seeded into 96-well plates at 4000 cells per well, and the proliferation activity of cells was detected on days 0, 1, 2, and 3, respectively. The CCK8 reagent (10 μl/ well) was added to the detection, and after incubation in the incubator for 4h, the optical density (OD) value of each well at a wavelength of 450 nm was measured by a microplate reader, and the cell proliferation curve was drawn.

### Cell migration and invasion assays

DU145 cells (5×10^4^) or HCT116 cells (1×10^5^) were suspended in 200 μL of RPMI 1640 or DMEM medium without serum and seeded into the upper transwell chambers, and then 600 μL of medium containing 20% FBS was added into the bottom chambers. Then, cells were cultured for 48 h to measure the migration and the invasion. Images were obtained and cells were counted with a microscope. And also, cell scratch healing assay to detect the migration ability of cells.

## Acknowledgements

We thank all study participants and the research staff of all parent studies for their contributions and commitment to this project. The data analyses were conducted using the Advanced Computing Center for Research and Education (ACCRE) at Vanderbilt University. This research is supported primarily by the grant from US National Institutes of Health R37 CA227130 and R01 CA269589 to X.G., R01 CA235553 and R01 CA202981 to W. Z., and Zhejiang provincial program for the Cultivation of High-level Innovative Health talents to W. L.. We also acknowledged the exceptional resources for consortia of colorectal (Supplementary Materials) and other cancer GWAS.

## Data Availability

Supplementary Table 1 provides the download information for the summary statistics of GWAS data for the six common cancers, including breast, ovary, prostate, colorectum, lung, and pancreas. Gene expression and alternative splicing data generated in normal tissues, were downloaded from GTEx consortium, and the individual-level genotype was downloaded from dbGaP (https://www.ncbi.nlm.nih.gov/projects/gap/cgi-bin/study.cgi?study_id=phs000424.v8.p2). Gencode annotation (v26.GRCh38) was downloaded from https://www.gencodegenes.org/human/release_26.html. For data of essentiality for proliferation and survival of cancer cells, we downloaded two comprehensive datasets including “sample_info.csv” and “Achilles_gene_effect.csv” from the DepMap portal. Remaining data sources and results are provided within the Article or Supplementary Tables.

## Code availability

The developed pipeline and main source R codes that are used in this work are available from the website of Xingyi Guo’s lab at Github: https://github.com/XingyiGuo/APA-WAS/.

## Author Contributions

X.G. and W.Z. conceptualized and designed the study. X.G., J. P., Y.Y., W.L. and W.Z., analyzed and interpreted the data and results. X. S., Y.Z., and W.L. performed the functional experiments. X.G., J.P., Y.Y., X.S., Q.C., G.G., R.P., G.R., P.V., A.P., S.G., G.C., U.P., J.L., and W.Z., acquired data. X.G., W.W., Z.C., R. T., G. J., J.H., Q.Z, and W.Z., provided administrative, technical, or material support. X.G., J.P., Y.Y., W.L. and W.Z., drafted the manuscript, with significant contributions from other authors. All authors critically revised and reviewed the manuscript.

## Supplementary Data

**Table S1:** A list of APA levels predicted by *cis-*genetic variants in normal multiple tissues of breast, ovary, prostate, colon transverse, lung and pancreas, at a prediction performance of R^2^ > 0.01 (10% correlation) at *P* < 0.05.

**Table S2.** GWAS summary statistics data of European descendants for breast, ovarian, colorectal, pancreatic, prostate, and lung cancers used in this study.

**Table S3:** Results of associations between genetically predicted APA levels and risk of breast, ovary, prostate, colorectal, lung and pancreatic cancer, at nominal *P* < 0.05.

**Table S4:** Breast cancer APA-WAS identified genes in this study reported in previous studies.

**Table S5:** Ovarian cancer APA-WAS identified genes in this study reported in previous studies.

**Table S6:** Prostate cancer APA-WAS identified genes in this study reported in previous studies.

**Table S7:** Colorectal cancer APA-WAS identified genes in this study reported in previous studies.

**Table S8:** Lung cancer APA-WAS identified genes in this study reported in previous studies.

**Table S9:** Colocalization analysis between 3’a QTL and GWAS signals for genes identified by the APA-WAS analysis.

**Table S10:** Joint TWAS analysis results based on multiple tissue models using MASHR and ACAT for genes identified by the APA-WAS analysis.

**Table S11:** Functional evidence of oncogenic roles for APA-WAS genes identified in breast, ovary, prostate, colorectal, and lung cancers.

**Table S12:** Results of 3’aQTL analyses for the lead variant included in a model of genetically predicted APA levels model for each of the APA-WAS-identified genes.

**Table S13:** A total of 33 APA-WAS identified genes supported by evidence of proximal regulation through putative regulatory 3’aQTL variants which are in strong LD (R^2^ >0.8) with the lead variant included in a model of genetically predicted APA level model.

